# Maternal prenatal immune activation associated with brain tissue microstructure and metabolite concentrations in newborn infants

**DOI:** 10.1101/2023.07.01.23292113

**Authors:** Marisa N. Spann, Ravi Bansal, Ezra Aydin, Angeliki Pollatou, Kiarra Alleyne, Margaret Bennett, Siddhant Sawardekar, Bin Cheng, Seonjoo Lee, Catherine Monk, Bradley S. Peterson

## Abstract

**Importance:** Few translational human studies have assessed the association of prenatal maternal immune activation with altered brain development and psychiatric risk in newborn offspring.

**Objective:** To identify the effects of maternal immune activation during the 2^nd^ and 3^rd^ trimesters of pregnancy on newborn brain metabolite concentrations, tissue microstructure, and longitudinal motor development.

**Design:** Prospective longitudinal cohort study conducted from 2012 – 2017.

**Setting:** Columbia University Irving Medical Center and Weill Cornell Medical College.

**Participants:** 76 nulliparous pregnant women, aged 14 to 19 years, were recruited in their 2^nd^ trimester, and their children were followed through 14 months of age.

**Exposure:** Maternal immune activation indexed by maternal interleukin-6 and C-reactive protein in the 2^nd^ and 3^rd^ trimesters of pregnancy.

**Main Outcomes and Measures:** The main outcomes included (1) newborn metabolite concentrations, measured as N-acetylaspartate, creatine, and choline using Magnetic Resonance Spectroscopy; (2) newborn fractional anisotropy and mean diffusivity measured using Diffusion Tensor Imaging; and (3) indices of motor development assessed prenatally and postnatally at ages 4- and 14-months.

**Results:** Maternal interleukin-6 and C-reactive protein levels in the 2^nd^ or 3^rd^ trimester were significantly positively associated with the N-acetylaspartate, creatine, and choline concentrations in the putamen, thalamus, insula, and anterior limb of the internal capsule. Maternal interleukin-6 was associated with fractional anisotropy in the putamen, insula, thalamus, precuneus, and caudate, and with mean diffusivity in the inferior parietal and middle temporal gyrus. C-reactive protein was associated with fractional anisotropy in the thalamus, insula, and putamen. Regional commonalities were found across imaging modalities, though the direction of the associations differed by immune marker. In addition, a significant positive association was observed between offspring motor development and both maternal interleukin-6 and C-reactive protein (in both trimesters) prenatally and 4- and 14-months of age.

**Conclusions and Relevance:** Using a healthy sample, these findings demonstrate that levels of maternal immune activation in mid- to late pregnancy associate with tissue characteristics in newborn brain regions primarily supporting motor integration/coordination and behavioral regulation and may lead to alterations in motor development.

**Key Points:** *Question:* What are the associations of prenatal maternal immune activation (MIA) with newborn brain microstructure, metabolite concentrations, and longitudinal motor development?

*Findings:* In this longitudinal cohort study we recruited 76 adolescent and young adult pregnant women and assessed maternal interleukin (IL)-6 and C-reactive protein (CRP) levels in the 2^nd^ and 3^rd^ trimesters. These pro-inflammatory markers were significantly associated with brain microstructure and metabolite concentrations in newborns, and longitudinal motor development (prenatally, 4- and 14-months of age).

*Meaning:* This study suggests that prenatal exposure to MIA has an influence on brain microstructure, metabolite concentration and motor development in offspring.

## INTRODUCTION

Maternal immune activation (MIA) refers to activation of the innate and adaptive immune system following infection, environmental or psychological stress, and chronic or acute physical illness. MIA is a process that is mediated by activation of inflammatory pathways resulting in increased levels of cytokines that cross the placental and blood-brain barriers^1^– it is the resulting molecular response that alters and shapes the neurodevelopment in the fetus^2–4^. Increasingly, evidence suggests MIA is an important environmental risk factor for neurodevelopmental brain dysfunction – with long-lasting consequences for offspring^5, 6^. Existing research (both human and non-human) has focused on long-term changes in brain development and postnatal behavioral outcomes. However, research examining both potential molecular and structural changes that occur in the fetal human brain and relating these potential changes to behavioral markers is limited.

Existing research into the influence of MIA on the developing human brain has largely focused on epidemiological data. These studies report associations between MIA during pregnancy and later neurodevelopment conditions (e.g., schizophrenia^3^). More recently human research studies have shifted to observing both structural and functional influences MIA can have on the developing brain. During activation the immune system releases several classes of proteins to stimulate an immune response. Two of the most commonly assayed immune biomarkers are interleukin (IL)-6, a pro-inflammatory cytokine, and/or C-reactive protein (CRP), an acute phase reactant^5, 7^, both used to assess the presence and severity of low-grade inflammation. Prior studies found prenatal exposure to IL-6 can influence newborn brain development – a period reflecting prenatal brain development ^3, 8^. For example, we found maternal IL-6 and CRP levels during the 3^rd^ trimester are associated with altered functional connectivity of the anterior cingulate and insula with brain regulatory regions, such as the medial prefrontal cortex, in newborns^9^. Others have demonstrated that MIA is associated with an increased strength of anatomical and functional associations across multiple newborn brain networks, including the salience, default mode, and frontoparietal networks^10, 11^, as well as reduced organization of the uncinate fasciculus, a white matter tract connecting the frontal cortex and amygdala^12^. These findings complement rodent and non-human primate studies suggesting that MIA exposure from immune-activating agents disrupt development of the hippocampus, prefrontal cortex, mid-temporal lobe, parietal lobe, insula, and cingulate cortex^13–15^. At a cellular level, these disruptions largely comprise reduced neuron growth, glial cell proliferation^16–23^ and altered cerebellar cytokine and synaptic protein expression^24, 25^ ultimately resulting in remodeling of the embryonic brain^26^.

Existing animal models also highlight potential differential influences of exposure on offspring brain and behavioral development. For example, MIA exposure in early pregnancy (first trimester) has been linked to accelerated increase in brain volume^26^ and greater deviations in postnatal neurodevelopmental trajectories in offspring, (e.g., sensorimotor gating and repetitive behaviors^27^). Conversely, MIA exposure later in pregnancy (second trimester) was not associated with behavioral impairments but related to altered developmental trajectories in emotional processing and reward regions of the brain – resulting in decreases in emotional processing ability in adulthood. This demonstrates that MIA during pregnancy can both disrupt and support development of neural systems, in turn this contributes to later offspring capacity for core areas essential for behavioral processes, such as self-regulation. Although pre-clinical findings suggest considerable effects of MIA on the offspring brain, those effects have not been extensively studied in humans^9–11^. Here we aim to provide further insight into low-grade inflammation in healthy pregnancies and its influence on offspring brain development, and in turn, later behavioral outcomes. Given the observed differential effects of MIA exposure *in utero* on the developing prenatal brain in animal models and existing research in this area – it is important to determine (1) whether these differential influences of MIA across gestation can be observed in humans, (2) how these differential brain effects may connect to behavior longitudinally from pre- to postnatal development.

One of the earliest brain structures to develop is the cerebellum, emerging from the roof of the rhombencephalon between 4-6 weeks post-conception^28^. This area of the brain is traditionally associated with motor movement and coordination; however, it also connects cortical and subcortical areas. It is via these connections the cerebellum acts as a modulator for many emotional and behavioural functions^29, 30^ including memory and language. Therefore, we would expect, in humans, to see the influence of low-grade inflammation from the prenatal stages influencing early brain development in offspring, observable via longitudinal behavioral outcomes, particularly in those behaviors associated with the cerebellum (i.e., motor movement).

To our knowledge, no studies have assessed the association of MIA with brain metabolite concentrations or tissue microstructure in newborns, which would shed light on the underlying molecular effects of MIA exposure in human newborns. Enhanced understanding of the influence of MIA on the developing brain and later behavioral outcomes will help inform future research and enable researchers and clinicals alike to observe potential deviations. For example, the current pandemic virus, Severe Acute Respiratory Syndrome Coronavirus 2 (SARS-CoV-2), demonstrated the link between prenatal infections and the cascading inflammatory risk to fetal development ^31–37^. This is the first prospective study to assess the association of MIA indices (circulating levels of inflammatory markers IL-6 and CRP) during pregnancy with magnetic resonance imaging (MRI) measures of microstructural organization and metabolite concentrations in the newborn brain. Given previous associations of MIA with altered newborn brain development^9, 11^. We hypothesized that MIA would be associated with increased microstructural organization, as measured by fractional anisotropy (FA) and mean diffusivity (ADC), and elevated concentrations of N-acetylaspartate (NAA), choline (Cho), and creatine (Cr) in prefrontal, temporo-parietal, cingulate, and basal ganglia regions. Furthermore, we hypothesize that these changes in offspring brain organization will be reflected in the longitudinal observations of motor development from pre- to postnatal life.

## METHODS

### Participants

Nulliparous adolescent and young women aged 14 to 19 years were recruited in the 2^nd^ trimester through the Departments of Obstetrics and Gynecology at Columbia University Irving Medical Center (CUIMC) and Weill Cornell Medical College, and through flyers posted in the CUIMC vicinity as part of a longitudinal study examining adolescent pregnancy behaviors and infant outcomes. Participants reported no major health problems and received routine prenatal care. Approval for the study was given by the Institutional Review Boards of the New York State Psychiatric Institute and of CUIMC. All mothers provided informed written consent. Exclusion criteria included use of recreational drugs, tobacco, alcohol, medications with an effect on cardiovascular function (e.g., beta blockers), or lacking English language fluency. Of the 324 adolescents enrolled during pregnancy, this report includes a sample of 76 infants from whom usable MRI data was obtained (*see supplemental Figure 1 for flow chart of study and data from each timepoint*).

### Study Procedures

#### Immune markers

During their 2^nd^ (24-27 weeks gestation) and 3^rd^ (34-37 weeks gestation) trimesters, women underwent phlebotomy to determine maternal IL-6 and CRP levels. IL-6 was measured using an enzyme-linked immunosorbent assay (ELISA) by R&D systems (Minneapolis, MN). The normal range values for healthy pregnant women during their first trimester is <3.52pg/ml and <4.40pg/ml second and third trimester^38^. CRP was measured using the Cobras Integra 400 Plus (Roche Diagnostics) turbid metric. Normal range values for this population are 0.4- 20.3mg/L during the second trimester and 0.4-8.1mg/L in the third^39^.

#### Electronic Health Records

Obstetrical and newborn electronic health records were used to gather information on the participants’ pregnancy, labor, and delivery.

#### Imaging techniques

Infants underwent an imaging protocol within the first weeks of postmenstrual life (mean 42.4, SD 1.6 weeks postmenstrual age at scan). Postmenstrual age (PMA) at the time of the scan was used to subsume variation in gestational age (GA) at birth and time since birth. Infants were assessed with both DTI and MRS. DTI measures both the direction and rate of diffusion water as influenced by tissue microstructure, utilizing both fractional anisotropy (FA) – indexing the degree to which water has a preferential direction of diffusion and average diffusion coefficient (ADC) – quantifying the overall directionless rate of the diffusion of water in each brain voxel. Infant scans were conducted during natural sleep with no sedation. Foam/wax ear plugs, and ear shields (Natus Medical Inc., San Carlos, CA) were used to damper scanner noise. MRI acquisition, MRS processing and DTI pulse sequence procedures are detailed in the Methods in the supplement.

#### Longitudinal motor development

Fetal movement (FM) data was collected via a single transabdominal doppler transducer using a Toitu MT 325 fetal echocardiograph (Toitu Co., Ltd, Tokyo, Japan). The Toitu system allows for capturing of fetal heart rate and movement. The received signal is processed through a series of filters, removing frequency components of the Doppler signal that are associated with both fetal and maternal somatic activities allowing for reliable distinction between the two^40^. Postnatal motor development was collected using the BSID-III at 4- and 14-months of age^41^. Scaled (age-standardized) scores were used in statistical analyses.

### Statistical Analyses

Analysis was conducted through SAS (Version 9.4) and R (Version 4.0.1). IL-6 and CRP were log-transformed due to their skewed distributions. Spearman correlations and ANOVA F-tests were used to assess the associations of MIA measures with continuous and categorical demographic variables, respectively. Spearman correlations were also used to assess the associations of MIA and brain measures with standardized scores for motor development (gross and fine) on the BSID-III. All continuous variables were standardized by subtracting their means and being divided by their standardized deviations. We applied multiple linear regression at each voxel in template space within each MRI modality to test our hypothesis that MIA would be associated with motor development in the newborn brain. For DTI analyses, the dependent measure was either FA or ADC, and for MPCSI analyses, it was NAA, Cr, or Cho concentration. The independent variable (MIA) was either prenatal CRP or IL-6. Covariates in all analyses were sex and infant PMA at the time of scan.

We used the topological false discovery rate (FDR) procedure^42^ at an FDR=0.05 to control for false positives when conducting multiple hypotheses testing across all voxels in the brain; p-values that survived the FDR correction were color-coded and mapped onto the T1- template. Furthermore, to account for the correlation in test statistics in MRI due to limited spatial resolution and data smoothing, we applied both a simple conservative modification of the FDR procedure. For additional post hoc analysis, see Methods in the Supplement.

## RESULTS

### Demographic characteristics

Maternal and newborn demographic characteristics are summarized in *Table 1*. The study included 76 infants (birthweight: M=3224.6g, SD=463.2, GA at birth: M=39.3, SD=1.3 weeks) and were scanned at an average of 42.4 (SD=1.6) weeks PMA. 63.5% of infants were male.

**Table 1.** Maternal and Newborn Demographics

| **Variables** | **n** | **Mean (SD)**<br>**or %** |
| --- | --- | --- |
| *Maternal* |  |  |
| Age at delivery, years | 63 | 18.2 (1.4) |
| Pre-pregnancy Body Mass Index (BMI) | 63 | 25.1 (6.2) |
| Years of Education |  |  |
| 8th grade | 1 | 1.6% |
| 9th grade | 7 | 11.1% |
| 10th grade | 8 | 12.7% |
| 11th grade | 22 | 34.9% |
| 12th grade or higher | 25 | 39.7% |
| Ethnicity |  |  |
| Not Hispanic/Latina | 6 | 9.5% |
| Hispanic/Latina | 57 | 90.5% |
| Type of Delivery |  |  |
| Vaginal spontaneous | 25 | 43.1% |
| Assisted vaginal | 23 | 39.7% |
| Emergent Cesarean section | 10 | 17.2% |
| Pregnancy Complications |  |  |
| None | 47 | 85.5% |
| Complications (infection) | 8 | 14.5% |
| *Newborn* |  |  |
| Gestational Age at Birth, wks | 63 | 39.3 (1.3) |
| Birth Weight, gms | 63 | 3224.6 (463.2) |
| Birth Head Circumference, cms | 52 | 33.9 (1.3) |
| Apgar 1 minute | 56 | 8.5 (1.2) |
| Apgar 5 minute | 56 | 8.5 (1.2) |
| Postmenstrual age at scan | 60 | 42.4 (1.6) |
| Sex |  |  |
| Female | 23 | 36.5% |
| Male | 40 | 63.5% |

#### Confounding variables

All participant characteristic variables (inclusive of maternal, newborn and pregnancy) and our primary variables of interest were analyzed to observe any significant relationships. Pregnancy complications (i.e., infections such as chorioamnionitis) were significantly associated with CRP during the 3^rd^ trimester (p=0.0003). In the newborn characteristics, there was a significant association between CRP during the 3^rd^ trimester and GA at time of birth (r=-0.34, p=0.04). No other significant associations were found between immune markers and demographic variables.

#### Maternal Immune Marker levels

IL-6 values during the second trimester, mean=1.3pg/ml (SD:0.7), and third mean=1.7pg/ml (SD:0.9). CRP in second trimester mean=5.3mg/L (SD:3.0) and mean=5.4mg/L in the third (SD:3.4). The correlation between IL-6 and CRP was not significant (R=0.39, p=0.08, DF=19)

### Diffusion Tensor Imaging

CRP in the 2^nd^ and 3^rd^ trimester was significantly and inversely associated with newborn FA values in the putamen, ALIC, and thalamus (p<.0001) (*top panel of Figure 1 and Supplemental Figure 2*); 2^nd^ trimester CRP levels were positively associated with ADC values in the orbital gyrus (p values varying between <.001 to <.05).

**Figure 1:**
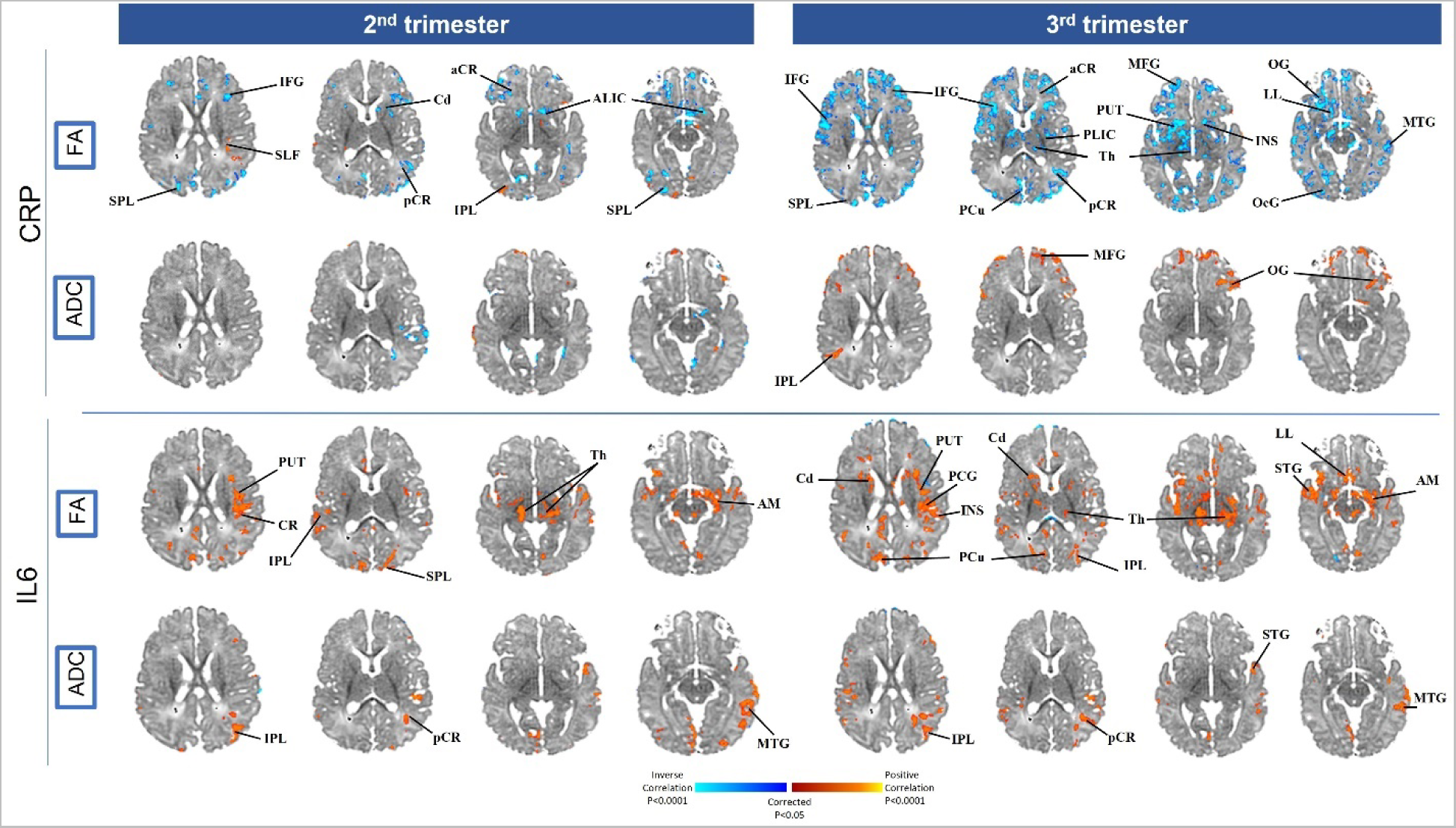
Association between diffusion tensor imaging (DTI), fractional anisotropy (FA) and mean diffusivity (ADC), and maternal immune activation through IL-6 and CRP during the 2^nd^ and 3^rd^ trimester. The red/yellow (positive) and purple/blue (inverse) areas show locations where maternal immune markers are associated with FA and ADC. aCR, anterior region of corona radiata; ALIC, Anterior limb of the internal capsule; Am, Amygdala; Cd, Caudate; CG, cingulum; CR, corona radiata; CC, corpus callosum; Cu, Cuneus; IFG, inferior frontal gyrus; IFO, inferior fronto-occipital; ITG, inferior temporal gyrus; Ins, Insula; IPL, inferior parietal lobule; LL, limbic lobe; MFG, medial frontal gyrus; MB, midbrain; MTG, middle temporal gyrus; OcG, occipital gyrus; OG, orbital gyrus; pCR, posterior region of corona radiata; PCG, precental gyrus; Pcu, Precuneus; PUT, Putamen; ST, stria terminalis; SFG, superior frontal gyrus; SFO, superior fronto-occipital; SLF, superior longitudinal fasciculus; SCR, superior region of corona radiata; SPL, superior parietal lobule; STG, superior temporal gyrus; Th, Thalamus; PLIC, posterior limb internal capsule.

IL-6 in the 2^nd^ and 3^rd^ trimester was positively associated with FA in the thalamus, insula, and putamen (p<.0001). In addition, 3^rd^ trimester IL-6 was positively associated with FA in the caudate and precuneus (p<.0001). Lastly, IL-6 also demonstrated significant positive correlation with ADC in both the 2^nd^ and 3^rd^ trimester in the inferior parietal and middle temporal gyrus (p values varying between <.001 to <.05) (*see bottom panel of Figure 1 and Supplemental Figure 2*).

### Magnetic Resonance Spectroscopy

CRP in the 2^nd^ and 3^rd^ trimesters was positively associated with newborn Cho, Cr, and NAA concentrations in regions identified using DTI, specifically the thalamus, insula, caudate, and precuneus (*Figure 2 and Supplemental Figure 3*). CRP during the 3^rd^ trimester was also positively associated with metabolite concentration in the anterior and posterior limb of the internal capsule (ALIC, PLIC) (*Figure 2 and Supplemental Figure 3*).

**Figure 2:**
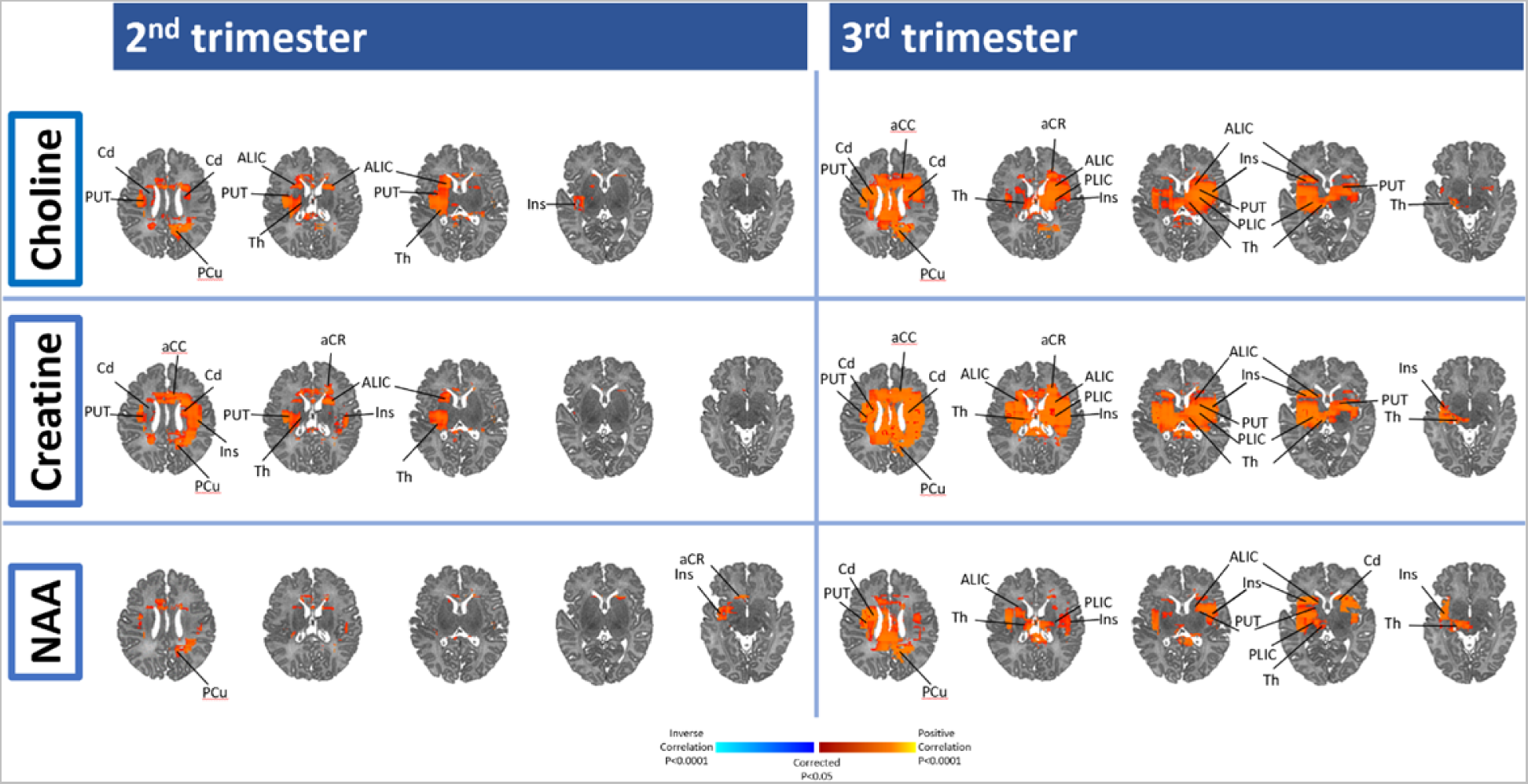
2^nd^ and 3^rd^ trimester maternal immune marker (CRP) association with neonatal brain metabolites. Using the whole brain, MRS, red/yellow (positive) and purple/blue (inverse) areas show locations where maternal immune markers are associated with Choline, Creatine, and *N*-acetylaspartate. aCC, anterior cingulate gyrus; ALIC indicates anterior limb internal capsule; aCR, anterior region of corona radiata; Cd, Caudate; CR, corona radiata; CC, corpus callosum; Cu, Cuneus; Ins, Insula; MFG, medial frontal gyrus; MB, midbrain; MTG, middle temporal gyrus; pCR, posterior region of corona radiata; PCu, Precuneus; PUT, Putamen; SFG : superior frontal gyrus, STG, superior temporal gyrus; Th, Thalamus; PLIC, posterior limb internal capsule.

IL-6 in the 2nd and 3rd trimesters was inversely associated with newborn Cho, Cr, and NAA in several regions, with additional inverse associations in the 2^nd^ trimester seen in the thalamus, putamen, insula and ALIC. IL-6 in the 3^rd^ trimester was inversely associated with Chlo and Cr in the thalamus, and with Cr in the ALIC and putamen (*Figure 3 and scatterplots in Supplemental Figure 3*), and positively with NAA in the putamen and PLIC.

**Figure 3:**
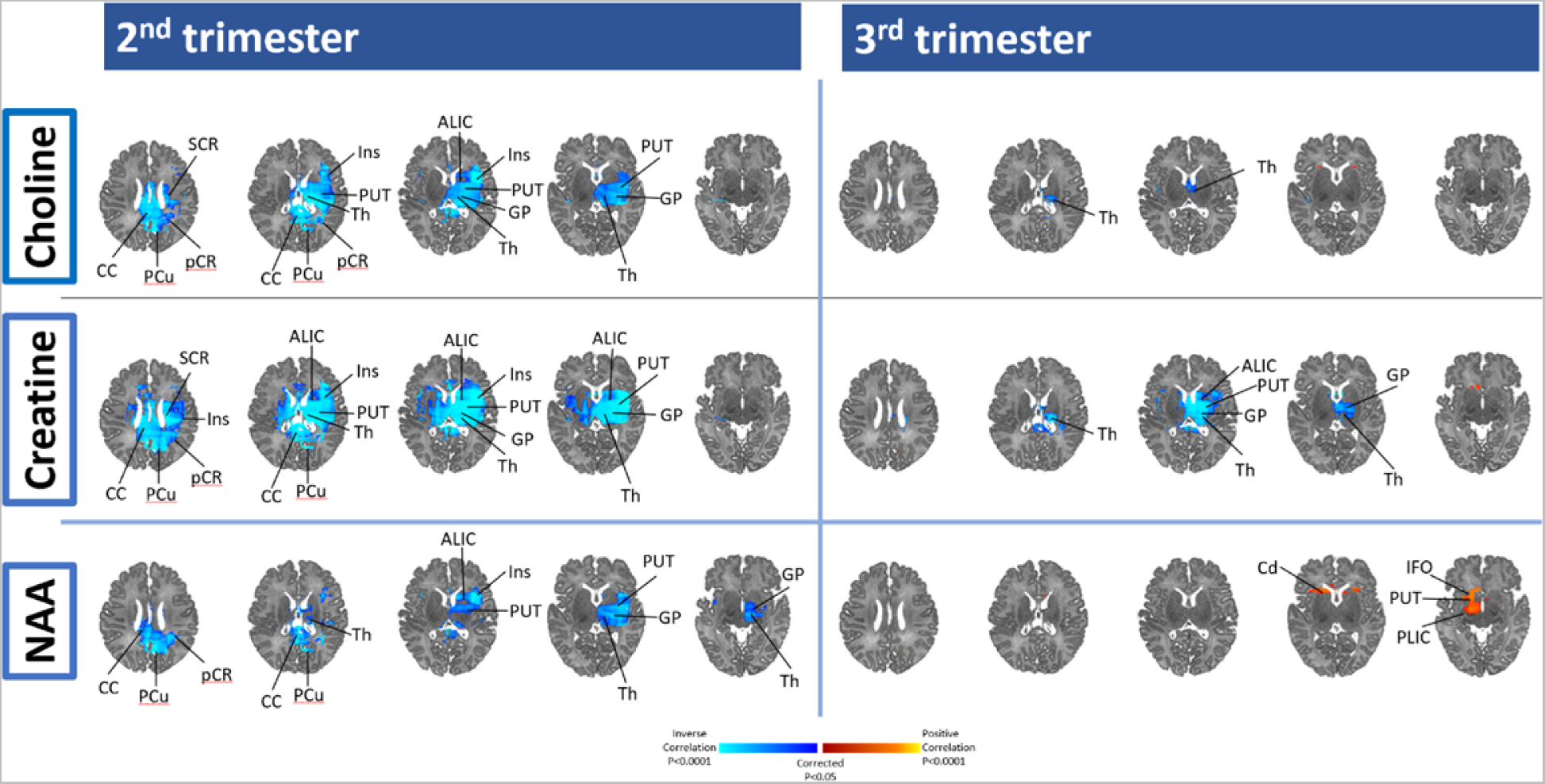
2^nd^ and 3^rd^ trimester maternal immune marker (IL-6) association with Neonatal brain metabolites. Using the whole brain, MRS, red/yellow (positive) and purple/blue (inverse) areas show locations where maternal immune markers are associated with Choline, Creatine, and *N*-acetylaspartate. ALIC indicates anterior limb internal capsule; aCR, anterior region of corona radiata; Cd, Caudate; CC: cingulate cortex; CR, corona radiata; CC, corpus callosum; Cu, Cuneus; GP, Globus Pallidus; IFO, inferior fronto-occipital; ITG, inferior temporal gyrus; Ins, Insula; MFG, medial frontal gyrus; MB, midbrain; MTG, middle temporal gyrus; pCR, posterior region of corona radiata; PUT, Putamen; SFG, superior frontal gyrus; SFO, superior fronto- occipital; SLF, superior longitudinal fasciculus; SCR, superior region of corona radiata; STG, superior temporal gyrus; Th, Thalamus; PLIC, posterior limb internal capsule.

### Comparison of Associations in 2^nd^ and 3^rd^ Trimesters

When we compared the strength of association between the immune markers and brain microstructural organization and biochemical levels (*see Supplemental Figure 4*), we found that the strength of associations was generally greater with 3^rd^ trimester immune markers and FA, and 2^nd^ trimester immune markers and brain metabolite levels. The differences across the two trimesters for the strength of association of immune markers with MRI measures were generally small (<0.2) (*Supplemental Figure 4 for values*).

### Associations of MIA with Longitudinal Motor Development

#### Prenatal motor development

During the 3^rd^ trimester both IL-6 (R=0.34, p=0.03) and CRP (R=0.41, p=0.01) showed a positive correlation with fetal movement. In the 2^nd^ trimester a significant association was only observed with CRP (R=0.41, p=0.01).

#### Postnatal motor development

IL-6 during the 2^nd^ trimester was significantly and positively associated with 4-month gross motor scores (R=0.34, p<0.04) and 14-month fine motor movement (R=0.48, p<0.00). IL-6 in the 3^rd^ trimester was positively associated with both fine motor (R=0.49, p<0.003) and gross motor scores (R=-0.37, p=0.03) at 14-months. CRP during the 2^nd^ trimester was significantly associated with fine motor scores (R=0.35, p=0.03) and overall (composite of fine and gross) motor scores (R=0.33, p=0.04) at 4-months of age.

### Mediation Analyses

The left thalamus significantly mediated the effect of maternal immune activation (maternal 2^nd^ trimester IL-6) on gross motor movement at 4-months of age (NIE=0.517, p = 0.021). The NDE was estimated as −0.290 (95% bootstrap confidence interval: [-0.896, 0.526]) and the NIE was estimated as 0.517 (95% bootstrap confidence interval: [0.172, 1.344]), and the percentage of effect mediated was 228% of the mediation. In addition, the left PLIC mediated the effect of maternal immune activation (maternal 2^nd^ trimester IL-6) on fine motor movement at 14-months of age (NIE=0.795, p = 0.036). The NIE was estimated as 0.220 (95% bootstrap confidence interval: [-0.667, 2.280]) and the NIE was estimated as 0.795 (95% bootstrap confidence interval: [-0.225, 2.093]), and the percentage of effect mediated was 78.3% of the mediation (*see Figure 4*). No other significant mediations were observed.

**Figure 4:**
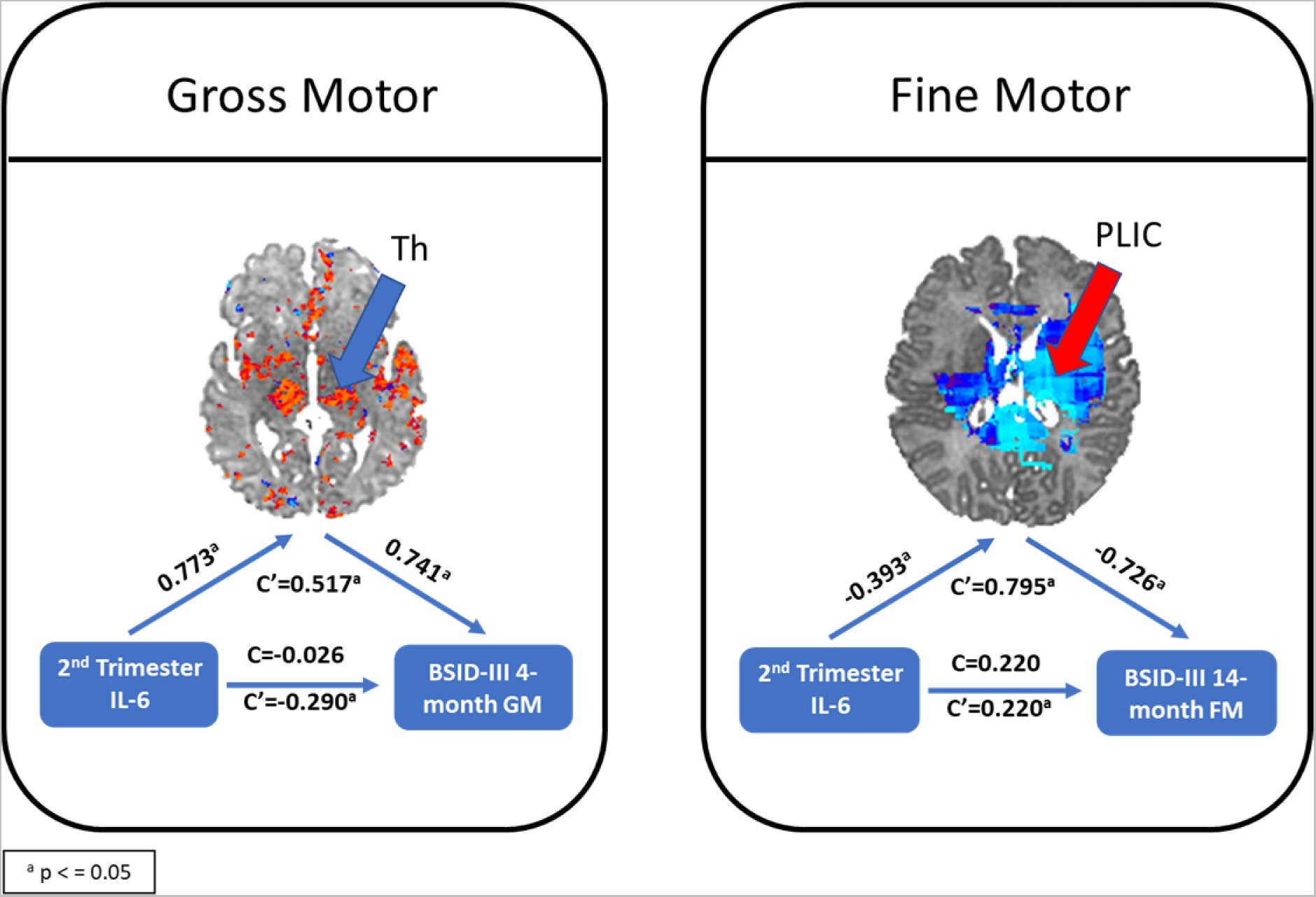
Mediation Model with 2^nd^ trimester IL-6, infant connectivity and BSID-III indices. *Left panel:* mediating effect of 2^nd^ trimester IL-6 to gross motor movement at 4-months of age through the thalamus. *Right panel:* mediating effect of 2^nd^ trimester IL-6 to fine motor movement at 14-months of age through the posterior limb internal capsule. These two data points had significant mediation in our study, but no other significant mediation was observed. C is the direct effect of the exposure on the outcome controlling for the mediator, while C’ the mediation effect, that the exposure changed the outcome through the mediator.

## DISCUSSION

In this prospective longitudinal study, we detected significant associations of prenatal MIA (indexed using CRP and IL-6) with microstructural measures of newborn brain tissue (FA and ADC) and metabolite concentrations (NAA, Cr, and Cho). These associations were generally consistent across the 2^nd^ and 3^rd^ trimesters, with the direction of associations differing with the specific MIA exposure measure and were centered on subcortical gray matter regions (e.g., thalamus, putamen). In addition, a significant association was observed between prenatal MIA (CRP and IL-6) and offspring motor behaviors. CRP concentration in the 2^nd^ trimester and both IL-6 and CRP concentrations during the 3^rd^ trimester were significantly associated with prenatal movement. Similarly, an association between offspring motor movement at 4- and 14-months of age was significantly associated with both immune markers during both 2^nd^ and 3^rd^ trimesters. Thus, demonstrating maternal inflammation during pregnancy (indexed using CRP and IL-6 concentrations) appears to have implications for the developing newborn brain that is reflected in observations of pre- and postnatal motor development.

The pathogenic importance of the timing of MIA exposure for brain development has been well documented in preclinical studies but has not been well characterized in human studies. Our findings suggest MIA during both 2^nd^ and 3^rd^ trimesters have comparable strength of associations with newborn brain measures and in similar regions across all modalities. During the 2^nd^ and 3^rd^ trimesters of pregnancy the fetal brain experiences rapid growth characterized by dendritic arborization, synaptogenesis and glial cell proliferation^43^. The observed significant inverse association between CRP and FA values putamen, insula, and thalamus during both trimesters, and a positive association of CRP with ADC values in the orbital gyrus during only the 2^nd^ trimester, are suggestive of reduced neuronal integrity and connectivity in the context of low-grade inflammation. Overall demonstrating regardless of the degree, immune activation during pregnancy, has an influence on the developing fetal brain.

In addition, we demonstrate a significant association between MIA markers and newborn microstructural indices and metabolite concentrations, primarily in subcortical brain regions linked to the cortico-striato-thalamic-cortical (CSTC) circuit, including the insula and precuneus. These findings compliment those from extensive rodent and non-human primate studies, and limited human studies, suggesting that prenatal MIA disrupts the development of widespread brain regions that include the hippocampus, prefrontal cortex, mid-temporal lobe, parietal lobe, insula, and cingulate cortex^13–15^. To date, one other study has considered newborn brain microstructure, also identifying an association with maternal IL-6, however this study was limited to only a single tract, the uncinate fasciculus^12^, and to our knowledge, none have considered brain metabolites.

In adults, the CSTC pathway controls important physiological functions, such as control of movement execution, and is a major site of synaptic dysfunction related to behavioral reinforcement and reward^44^. NAA is present primarily in the mitochondria of neurons and therefore is considered a good surrogate marker for neuronal health^45^. It contributes to signaling between neurons and oligodendrocytes and participates in myelin synthesis by oligodendrocytes^46^. Cho is involved in membrane synthesis and degradation, and therefore used as a marker of the structural integrity and turnover of cell membranes^47^. Lastly Cr is mainly involved in the storage and transfer of energy in metabolically active tissues (e.g., the brain). The observed CRP related elevations in NAA, Cho and Cr suggest the presence of greater neuronal density and higher cell membrane turnover in the cingulate gray matter and associated white matter pathways of CSTC circuits. Conversely, in IL-6, inverse associations (at both trimesters) were observed in relation to NAA, Cho and Cr concentrations suggesting a degradation of neuronal density and cell membrane turnover in these areas of the CSTC circuit. These contrasting findings suggest there may be counter effects of different immune markers, some being protective, supporting the developing fetal brain and others being disruptive, negatively impacting the early formation of neuronal pathways and structural growth. However, further studies are necessary with more extensive immune profiles to further elucidate these early influences. While further study is needed to distinguish between these differing directions of association and the implications – our findings suggest that both immune markers (CRP and IL- 6) play an important role in supporting the early development of the fetal brain. These results demonstrate a relationship between low-grade inflammation and the developing fetal brain in uncomplicated, healthy pregnancies, supporting the need to further explore this interplay between the maternal immune environment and the fetal brain.

Lastly, many of the regions implicated within this study have structural and functional connections with one another and support coordinated higher-order cognition, motor functions and behavioral regulation. For example, basal ganglia, thalamus, and insula are involved in motor coordination, cognitive control, and emotional processing^48^. The anterior and posterior limbs of the internal capsule have short- and long-range fibers that interconnect widespread cortical regions with the basal ganglia, thalamus, and brainstem^49^. Prior studies have reported that prenatal MIA is associated with offspring behavioral disturbances, including increased behavioral reactivity or disinhibition, and deficits in emotion regulation, attention, cognition and memory^11, 12, 50–54^. Our findings uniquely contribute to the existing literature by demonstrating the longitudinal associations between MIA and motor development from *in utero*. Results highlight the potential associations between MIA markers, prenatal motor movement, newborn CSTC circuit connectivity and later infant motor coordination. Furthermore, we saw the additional influence of MIA to several brain regions consistent with their roles in motor coordination - the basal ganglia, thalamus, and ALIC^12, 55^. Our results are consistent with those of a recent study that reported maternal IL-6 at 26 weeks gestation in HIV-exposed but uninfected toddlers was associated with poorer motor skills at 24 to 28 months of age^55^. IL-6 is associated with a lower number of inhibitory synapses and altered morphology of dendritic spines^56^. In contrast, CRP influences synaptic pruning, among other brain development processes.^57^ Therefore, alterations maternal CRP or IL-6 during pregnancy may lead to deviations in neuronal development and reduced connectivity across short and long-range brain networks, leading to these observed disruptions in longitudinal motor development.

### Strengths and Limitations

To our knowledge, this is one of the few infant neuroimaging studies in humans to consider associations with more than one prenatal immune marker and longitudinal motor development, prior to postnatal influences. Interestingly, while directionality of associations sometimes differed between the immune markers, we saw consistency in influence of these markers (CRP and IL-6) across brain regions. In particular in those involved in CSTC pathways. It is also important to note this study has several limitations.

Challenges with follow-up assessments through toddler age led to a reduced sample with behavioral outcome data (*see Figure 1*), limiting statistical power to assess the potential brain-based mediation of associations of MIA with infant motor outcomes. As the original study was interested in stress and nutrition in the context of adolescent pregnancy, the majority of participants were adolescent women and of Hispanic descent. As such, the results may not be generalizable to a more diverse population. Our sample had very few major maternal infections during pregnancy, as reflected in the low rate of obstetrical complications. While it is possible that our measures of MIA reflect inflammation due to infection or other physical health conditions, the findings more likely reflect normal variation of immune levels in the context of adolescent pregnancy and minor infectious illnesses during pregnancy, thus only assessing low level inflammation. Results in the context of significant MIA may differ. Lastly, there was a limited sample of cortical mantel due to the need for saturation bands that overlap portions of the cortex and the low number of directions for DTI due to our relatively young (newborn) sample.

### Conclusion

Given neuroinflammation is consistently identified as playing a role in multiple neuropsychiatric and neurodevelopmental conditions it is important for researchers and clinicians alike to understand the prenatal influence on the next generation from low-grade inflammation during uncomplicated, healthy pregnancies. Future studies with MIA measures that can isolate the time course of innate and adaptive immune pathways could have great utility in establishing whether metabolic (e.g., inflammatory cytokines) or environmental (e.g., microbial, particulate matter) stressors are the primary drivers of the effects of MIA on early brain development.

## Data Availability

All data produced in the present study can be made available upon reasonable request to the authors

## Acknowledgements

We wish to thank the women who participated in this study, our research assistants and staff, Alida Davis, Ashley Rainford, Grace Liu, Mei Ju Chen and Kirwan Walsh for dedicated help with participant engagement and data collection.

## References

1. Mueller FS, Scarborough J, Schalbetter SM, et al. Behavioral, neuroanatomical, and molecular correlates of resilience and susceptibility to maternal immune activation. Mol Psychiatry. 02 2021;26(2):396–410. doi:10.1038/s41380-020-00952-8

2. Ponzio NM, Servatius R, Beck K, Marzouk A, Kreider T. Cytokine levels during pregnancy influence immunological profiles and neurobehavioral patterns of the offspring. Ann N Y Acad Sci. Jun 2007;1107:118–28. doi:10.1196/annals.1381.013

3. Purves-Tyson TD, Weber-Stadlbauer U, Richetto J, et al. Increased levels of midbrain immune-related transcripts in schizophrenia and in murine offspring after maternal immune activation. Mol Psychiatry. Mar 2021;26(3):849–863. doi:10.1038/s41380-019-0434-0

4. Machado CJ, Whitaker AM, Smith SE, Patterson PH, Bauman MD. Maternal immune activation in nonhuman primates alters social attention in juvenile offspring. Biol Psychiatry. May 01 2015;77(9):823–32. doi:10.1016/j.biopsych.2014.07.035

5. Estes ML, McAllister AK. Maternal immune activation: Implications for neuropsychiatric disorders. Science. Aug 19 2016;353(6301):772–7. doi:10.1126/science.aag3194

6. Monk C, Spicer J, Champagne FA. Linking prenatal maternal adversity to developmental outcomes in infants: the role of epigenetic pathways. Dev Psychopathol. Nov 2012;24(4):1361–76. doi:10.1017/S0954579412000764

7. Hunter CA, Jones SA. IL-6 as a keystone cytokine in health and disease. Nat Immunol. May 2015;16(5):448–57. doi:10.1038/ni.3153

8. Rasmussen JM, Graham AM, Gyllenhammer LE, et al. Neuroanatomical Correlates Underlying the Association Between Maternal Interleukin 6 Concentration During Pregnancy and Offspring Fluid Reasoning Performance in Early Childhood. Biol Psychiatry Cogn Neurosci Neuroimaging. 01 2022;7(1):24–33. doi:10.1016/j.bpsc.2021.03.007

9. Spann MN, Monk C, Scheinost D, Peterson BS. Maternal Immune Activation During the Third Trimester Is Associated with Neonatal Functional Connectivity of the Salience Network and Fetal to Toddler Behavior. J Neurosci. Mar 2018;38(11):2877–2886. doi:10.1523/JNEUROSCI.2272-17.2018

10. Graham AM, Rasmussen JM, Rudolph MD, et al. Maternal Systemic Interleukin-6 During Pregnancy Is Associated With Newborn Amygdala Phenotypes and Subsequent Behavior at 2 Years of Age. Biol Psychiatry. Jan 2018;83(2):109–119. doi:10.1016/j.biopsych.2017.05.027

11. Rudolph MD, Graham AM, Feczko E, et al. Maternal IL-6 during pregnancy can be estimated from newborn brain connectivity and predicts future working memory in offspring. Nat Neurosci. May 2018;21(5):765–772. doi:10.1038/s41593-018-0128-y

12. Rasmussen JM, Graham AM, Entringer S, et al. Maternal Interleukin-6 concentration during pregnancy is associated with variation in frontolimbic white matter and cognitive development in early life. Neuroimage. 01 2019;185:825–835. doi:10.1016/j.neuroimage.2018.04.020

13. Short SJ, Lubach GR, Karasin AI, et al. Maternal influenza infection during pregnancy impacts postnatal brain development in the rhesus monkey. Biol Psychiatry. May 2010;67(10):965–73. doi:10.1016/j.biopsych.2009.11.026

14. Bland ST, Beckley JT, Young S, et al. Enduring consequences of early-life infection on glial and neural cell genesis within cognitive regions of the brain. Brain Behav Immun. Mar 2010;24(3):329–38. doi:10.1016/j.bbi.2009.09.012

15. Patterson PH. Maternal infection: window on neuroimmune interactions in fetal brain development and mental illness. Curr Opin Neurobiol. Feb 2002;12(1):115–8.

16. Brown AS, Hooton J, Schaefer CA, et al. Elevated maternal interleukin-8 levels and risk of schizophrenia in adult offspring. Am J Psychiatry. May 2004;161(5):889–95.

17. Brown AS, Derkits EJ. Prenatal infection and schizophrenia: a review of epidemiologic and translational studies. Am J Psychiatry. Mar 2010;167(3):261–80. doi:10.1176/appi.ajp.2009.09030361

18. Bilbo SD. How cytokines leave their mark: the role of the placenta in developmental programming of brain and behavior. Brain Behav Immun. May 2011;25(4):602–3. doi:10.1016/j.bbi.2011.01.018

19. Richetto J, Calabrese F, Meyer U, Riva MA. Prenatal versus postnatal maternal factors in the development of infection-induced working memory impairments in mice. Brain Behav Immun. Oct 2013;33:190–200. doi:10.1016/j.bbi.2013.07.006

20. Gilmore JH, Jarskog LF, Vadlamudi S. Maternal infection regulates BDNF and NGF expression in fetal and neonatal brain and maternal-fetal unit of the rat. J Neuroimmunol. May 2003;138(1-2):49–55.

21. Bilbo SD, Barrientos RM, Eads AS, et al. Early-life infection leads to altered BDNF and IL-1beta mRNA expression in rat hippocampus following learning in adulthood. Brain Behav Immun. May 2008;22(4):451–5. doi:10.1016/j.bbi.2007.10.003

22. Marx CE, Vance BJ, Jarskog LF, Chescheir NC, Gilmore JH. Nerve growth factor, brain-derived neurotrophic factor, and neurotrophin-3 levels in human amniotic fluid. Am J Obstet Gynecol. Nov 1999;181(5 Pt 1):1225–30.

23. Gilmore JH, Jarskog LF, Vadlamudi S. Maternal poly I:C exposure during pregnancy regulates TNF alpha, BDNF, and NGF expression in neonatal brain and the maternal-fetal unit of the rat. J Neuroimmunol. Feb 2005;159(1-2):106–12. doi:10.1016/j.jneuroim.2004.10.008

24. Aavani T, Rana SA, Hawkes R, Pittman QJ. Maternal immune activation produces cerebellar hyperplasia and alterations in motor and social behaviors in male and female mice. Cerebellum. Oct 2015;14(5):491–505. doi:10.1007/s12311-015-0669-5

25. Pendyala G, Chou S, Jung Y, et al. Maternal Immune Activation Causes Behavioral Impairments and Altered Cerebellar Cytokine and Synaptic Protein Expression. Neuropsychopharmacology. Jun 2017;42(7):1435–1446. doi:10.1038/npp.2017.7

26. Guma E, Bordeleau M, González Ibáñez F, et al. Differential effects of early or late exposure to prenatal maternal immune activation on mouse embryonic neurodevelopment. Proc Natl Acad Sci U S A. Mar 22 2022;119(12):e2114545119. doi:10.1073/pnas.2114545119

27. Guma E, Bordignon PDC, Devenyi GA, et al. Early or Late Gestational Exposure to Maternal Immune Activation Alters Neurodevelopmental Trajectories in Mice: An Integrated Neuroimaging, Behavioral, and Transcriptional Study. Biol Psychiatry. 09 01 2021;90(5):328–341. doi:10.1016/j.biopsych.2021.03.017

28. Garel C, Fallet-Bianco C, Guibaud L. The fetal cerebellum: development and common malformations. J Child Neurol. Dec 2011;26(12):1483–92. doi:10.1177/0883073811420148

29. Schmahmann JD, Sherman JC. Cerebellar cognitive affective syndrome. Int Rev Neurobiol. 1997;41:433–40. doi:10.1016/s0074-7742(08)60363-3

30. Schmahmann JD, Pandya DN. The cerebrocerebellar system. Int Rev Neurobiol. 1997;41:31–60. doi:10.1016/s0074-7742(08)60346-3

31. Schwartz DA, Dhaliwal A. infections in pregnancy with covid-19 and other respiratory rna virus diseases are rarely, if ever, transmitted to the fetus: experiences with coronaviruses, hpiv, hmpv rsv, and influenza. Arch Pathol Lab Med. Apr 2020;doi:10.5858/arpa.2020-0211-SA

32. Penfield CA, Brubaker SG, Limaye MA, et al. Detection of SARS-COV-2 in Placental and Fetal Membrane Samples. Am J Obstet Gynecol MFM. May 2020:100133. doi:10.1016/j.ajogmf.2020.100133

33. Martins-Filho PR, Tanajura DM, Santos HP, Santos VS. COVID-19 during pregnancy: Potential risk for neurodevelopmental disorders in neonates? Eur J Obstet Gynecol Reprod Biol. May 2020;doi:10.1016/j.ejogrb.2020.05.015

34. Li R, Yin T, Fang F, et al. Potential risks of SARS-CoV-2 infection on reproductive health. Reprod Biomed Online. 07 2020;41(1):89–95. doi:10.1016/j.rbmo.2020.04.018

35. Di Mascio D, Khalil A, Saccone G, et al. Outcome of Coronavirus spectrum infections (SARS, MERS, COVID 1-19) during pregnancy: a systematic review and meta-analysis. Am J Obstet Gynecol MFM. Mar 2020:100107. doi:10.1016/j.ajogmf.2020.100107

36. Rogers JP, Chesney E, Oliver D, et al. Psychiatric and neuropsychiatric presentations associated with severe coronavirus infections: a systematic review and meta-analysis with comparison to the COVID-19 pandemic. Lancet Psychiatry. May 2020;doi:10.1016/S2215-0366(20)30203-0

37. Iqbal A, Burrin C, Aydin E, Beardsall K, Wong H, Austin T. Generation COVID-19 - Should the foetus be worried? Acta Paediatr. Mar 2021;110(3):759–764. doi:10.1111/apa.15693

38. Fu Y, Tang L, Hu M, Xiang Z, Hu Y. Changes of serum interleukin-6 in healthy pregnant women and establishment of relevant reference intervals. Clin Chim Acta. Mar 2020;502:116–119. doi:10.1016/j.cca.2019.12.013

39. Abbassi-Ghanavati M, Greer LG, Cunningham FG. Pregnancy and laboratory studies: a reference table for clinicians. Obstet Gynecol. Dec 2009;114(6):1326–1331. doi:10.1097/AOG.0b013e3181c2bde8

40. Doyle C, Werner E, Feng T, et al. Pregnancy distress gets under fetal skin: Maternal ambulatory assessment & sex differences in prenatal development. Dev Psychobiol. Jul 2015;57(5):607–25. doi:10.1002/dev.21317

41. Bayley N. Scales of Infant and Toddler Development-Third Edition: Administration Manual. Harcourt Assesment; 2005.

42. Chumbley J, Worsley K, Flandin G, Friston K. Topological FDR for neuroimaging. Neuroimage. Feb 15 2010;49(4):3057–64. doi:10.1016/j.neuroimage.2009.10.090

43. Tau GZ, Peterson BS. Normal development of brain circuits. Neuropsychopharmacology. Jan 2010;35(1):147–68. doi:10.1038/npp.2009.115

44. Graybiel AM, Aosaki T, Flaherty AW, Kimura M. The basal ganglia and adaptive motor control. Science. Sep 23 1994;265(5180):1826–31. doi:10.1126/science.8091209

45. Zhu H, Barker PB. MR spectroscopy and spectroscopic imaging of the brain. Methods Mol Biol. 2011;711:203–26. doi:10.1007/978-1-61737-992-5_9

46. Moffett JR, Ross B, Arun P, Madhavarao CN, Namboodiri AM. N-Acetylaspartate in the CNS: from neurodiagnostics to neurobiology. Prog Neurobiol. Feb 2007;81(2):89–131. doi:10.1016/j.pneurobio.2006.12.003

47. Rae CD. A guide to the metabolic pathways and function of metabolites observed in human brain 1H magnetic resonance spectra. Neurochem Res. Jan 2014;39(1):1–36. doi:10.1007/s11064-013-1199-5

48. Uddin LQ. Salience processing and insular cortical function and dysfunction. Nat Rev Neurosci. 01 2015;16(1):55–61. doi:10.1038/nrn3857

49. Imperati D, Colcombe S, Kelly C, et al. Differential development of human brain white matter tracts. PLoS One. 2011;6(8):e23437. doi:10.1371/journal.pone.0023437

50. Bilbo SD, Biedenkapp JC, Der-Avakian A, Watkins LR, Rudy JW, Maier SF. Neonatal infection-induced memory impairment after lipopolysaccharide in adulthood is prevented via caspase-1 inhibition. J Neurosci. Aug 2005;25(35):8000–9. doi:10.1523/JNEUROSCI.1748-05.2005

51. Bilbo SD, Yirmiya R, Amat J, Paul ED, Watkins LR, Maier SF. Bacterial infection early in life protects against stressor-induced depressive-like symptoms in adult rats. Psychoneuroendocrinology. Apr 2008;33(3):261–9. doi:10.1016/j.psyneuen.2007.11.008

52. Malkova NV, Yu CZ, Hsiao EY, Moore MJ, Patterson PH. Maternal immune activation yields offspring displaying mouse versions of the three core symptoms of autism. Brain Behav Immun. May 2012;26(4):607–16. doi:10.1016/j.bbi.2012.01.011

53. Patterson PH. Immune involvement in schizophrenia and autism: etiology, pathology and animal models. Behav Brain Res. Dec 2009;204(2):313–21. doi:10.1016/j.bbr.2008.12.016

54. Ramsay H, Surcel HM, Björnholm L, Kerkelä M, Khandaker GM, Veijola J. Associations Between Maternal Prenatal C-Reactive Protein and Risk Factors for Psychosis in Adolescent Offspring: Findings From the Northern Finland Birth Cohort 1986. Schizophr Bull. 04 29 2021;47(3):766–775. doi:10.1093/schbul/sbaa152

55. Sevenoaks T, Wedderburn CJ, Donald KA, et al. Association of maternal and infant inflammation with neurodevelopment in HIV-exposed uninfected children in a South African birth cohort. Brain Behav Immun. 01 2021;91:65–73. doi:10.1016/j.bbi.2020.08.021

56. Wei H, Chadman KK, McCloskey DP, et al. Brain IL-6 elevation causes neuronal circuitry imbalances and mediates autism-like behaviors. Biochim Biophys Acta. Jun 2012;1822(6):831–42. doi:10.1016/j.bbadis.2012.01.011

57. Stephan AH, Barres BA, Stevens B. The complement system: an unexpected role in synaptic pruning during development and disease. Annu Rev Neurosci. 2012;35:369–89. doi:10.1146/annurev-neuro-061010-113810

