## Supplemental Materials for "Maternal prenatal immune activation associated with brain tissue microstructure and metabolite concentrations in newborn infants"

**eMethods**

**MRI Acquisition**

As previously described^39^, infants underwent MRI scanning on a 3T MR scanner using an 8-channel head coil. Multiplanar chemical shift imaging (MPCSI) data were acquired in six axial oblique slices parallel to the AC-PC line, with the second bottom-most slice containing the AC-PC plane. Parameters for the MPCSI sequence were TR=2800ms, TE=144ms, spectral width=2000Hz, number of complex data points=512, field of view=24cm, slice thickness=10.0mm, slice spacing=2.0mm, number of phase encoding steps=24 x 24. Water signal suppression was achieved using the CHESS sequence. Lipid signal was suppressed by placing eight angulated saturation bands around the brain. The MPCSI data were spatially registered to a template brain using a localizer image of high in-plane resolution in the same orientation and slice locations as the MPCSI data with TR=300ms, TE=10ms, FOV=24cm, slice thickness=10.0mm, spacing=2.0mm, matrix=256x128, image zero-padded to 256x256.

**MRS Processing**

MPCSI data were spatially normalized using the T1-weighted overlay image (i.e., the localizer image acquired) in nearly ideal alignment with the MPCSI data^40^~~.~~ As described previously^41^, we measured NAA, Chol, and Cr, values were normalized to the noise in the spectrum across participants. MPCSI data were preprocessed using the 3DiCSI software package (http://hatch.cpmc.columbia.edu/software.html), which identified MPCSI voxels inside the brain^40^. Spectra were fit for NAA, Cr, Chol using Gaussian-Lorentzian curves and least-squares. Areas under the curves estimated metabolite concentrations in each voxel. Quality control included inspection of each spectrum, rejecting spectra with lipid contamination, insufficient water suppression, unresolved Cr and Chol, or linewidth >12Hz. Background noise was also computed as the standard deviation of the part of the real spectrum free from metabolite signal. Finally, a spectroscopic image for each metabolite was generated as the ratio of peak area to background noise for each voxel, accounting for variations in receiver and transmitter gain.

The spectroscopic images were corrected for partial-voluming (variable gray- vs. white-matter content across MPCSI voxels) and for the MPCSI point-spread function (dispersion of the MR signal into neighboring voxels). Partial volume correction employed linear regression to estimate metabolite concentration in gray matter and white matter using the levels of that metabolite and the proportions of gray and white matter in neighboring MRS voxels. The proportions of gray and white matter were estimated by similarity transforming (three rotations and three translations) MPCSI data into each participant’s T1-weighted image, which was segmented to define gray and white matter. Lastly, partial-volume corrected metabolite images resampled into the high-resolution T1 during were spatial normalized to a template brain using high dimensional nonlinear transformation.

**DTI Pulse Sequence**

DTI slices were acquired in an axial oblique orientation parallel to the anterior–posterior commissure line using single-shot echo planar DTI imaging sequence, with repetition time=13,92 5ms, echo time=~74ms, field of view=19x19cm^2^, flip=90°, matrix=132x128 (acceleration factor=2) zero-padded to 256x256, for 60 oblique axial slices positioned parallel to the anterior–posterior commissure line, slices thickness=2.0 mm. We acquired 3 baseline images with *b*=0 s/mm2, and 11 diffusion-weighted images at *b*=600 s/mm^2^ with diffusion gradients applied in 11 directions sampling 3D space uniformly.

The diffusion tensor was computed at each voxel by fitting an ellipsoid to the diffusion-weighted imaging data acquired along 11 gradient directions and 3 baseline images. This was achieved using a Levenberg–Marquardt algorithm for a robust nonlinear least squares fit, while constraining the diffusion tensor to be positive definite^42^. FA maps generated from the diffusion tensor model were then spatially normalized to the template brain using a rigid body similarity transformation, followed by a nonlinear warping using a method based on fluid dynamics^43^.

**POST HOC STATISTICAL ANALYES**

**Mediation Analyses** We performed mediation analyses to investigate whether infant connectivity mediates the effect of MIA on motor development in the newborn brain. The mediation analysis was performed using the CAUSALMED procedure in SAS version 9.4 to estimate and test the significance of direct and indirect effects. The 95% bias corrected bootstrap confidence intervals for the natural direct effect (NDE) and natural indirect effect (NIE) were constructed based on 1000 bootstrap samples. For clear interpretation of the mediation effect, interaction between the maternal immune activation and the infant connectivity was not included in the model.

**Comparing MIA-Brain Correlations across Trimesters** Spearman correlations were calculated for MIA measures in each of the 2^nd^ and 3^rd^ trimesters with newborn DTI and MPCSI measures to determine whether similar patterns of association were observed across trimesters. We compared the correlations across the trimesters using bootstrapping with replacement. Correlation coefficients were calculated in each bootstrapped sample and then compared using paired t-tests with Bonferroni correction for multiple comparisons. Cohen’s d statistics were also calculated from the set of correlation coefficients as a measure of effect size across trimesters.

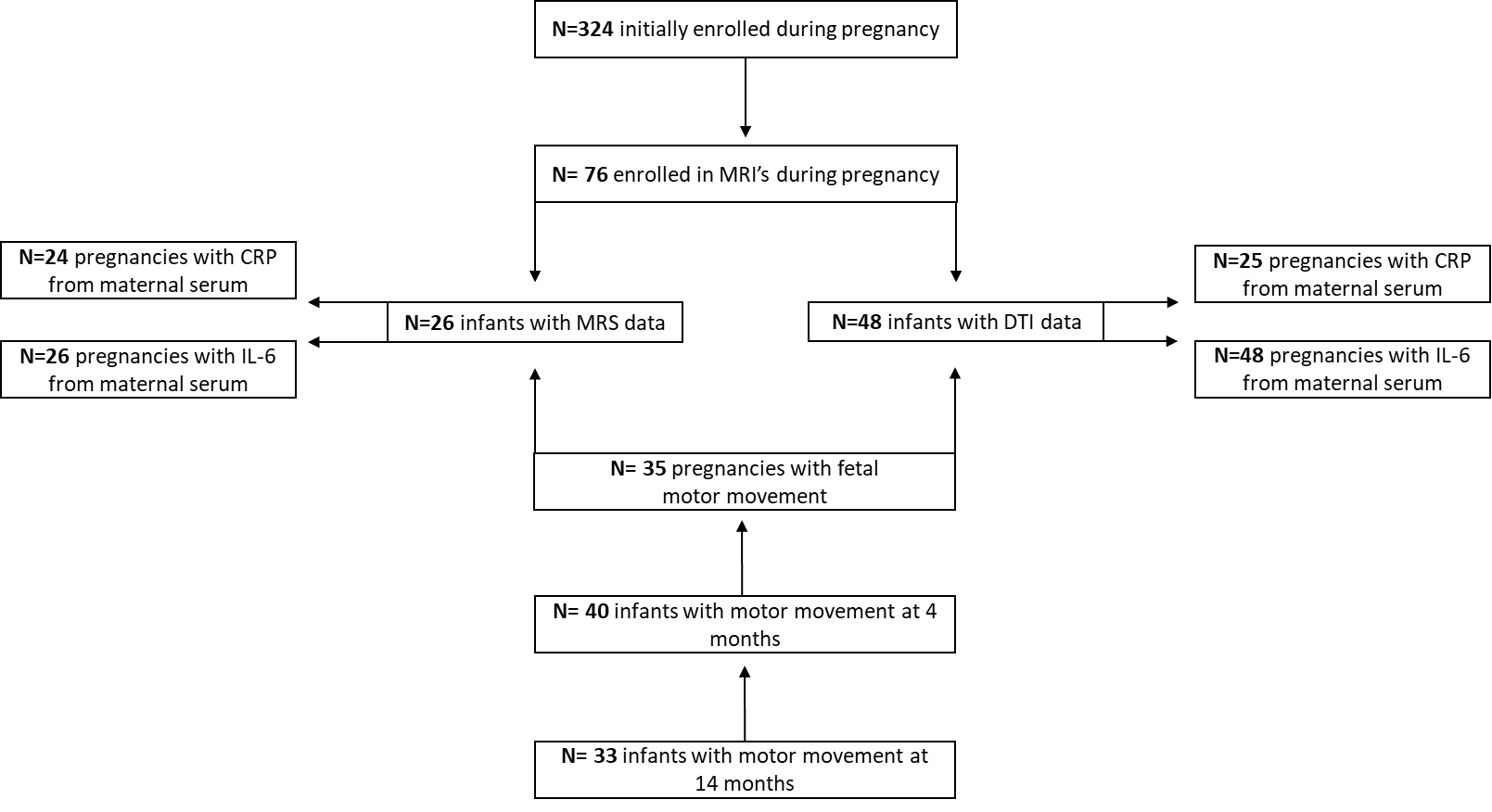

**Supplemental Figure 1: Flow chart of study.** Showing different cohort sizes for each comparison of maternal immune markers (IL-6 and CRP) to offspring outcome measures (neuroimaging; MRS and DTI and motor movement prenatally, 4- and 14-months).

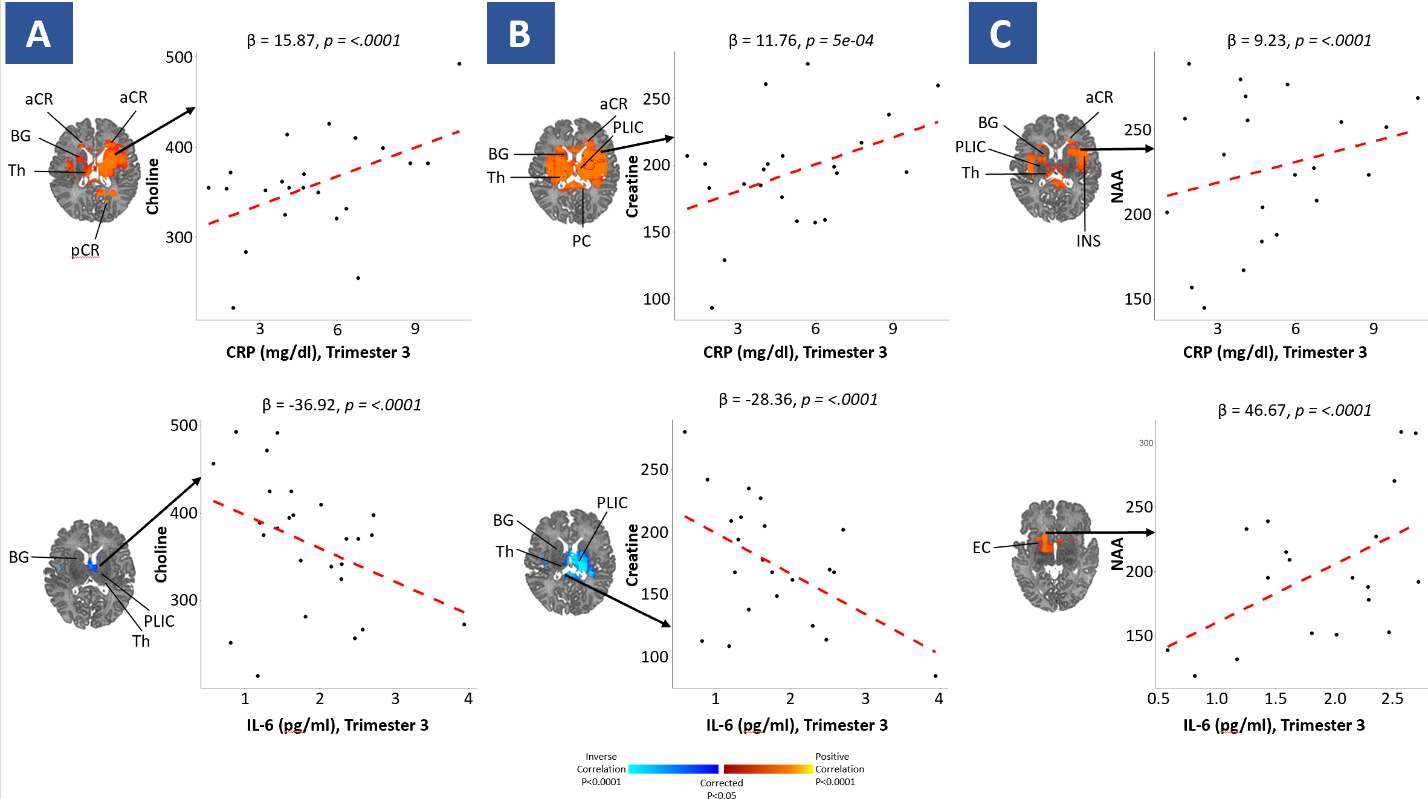

**Supplemental Figure 2:** **Parts A and B represent the association between DTI, FA, the dependent and maternal immune activation through IL-6 and CRP respectively during the 3^rd^ trimester as the independent, while part C represents the associations of 3^rd^ trimester IL-6 and CRP with Newborn NAA Concentrations**. The red/yellow (positive) and purple/blue (inverse) areas show locations where maternal immune markers are associated with each metabolite. Abbreviations: aCR, anterior corona radiata; pCR, posterior corona radiata; PLIC, posterior limb of the internal capsule; BG, basal ganglia; EC, external capsule; Th, thalamus; CC, cingulate cortex; Caud, Caudate nucleus; Ins, Insula; PUT, Putamen.

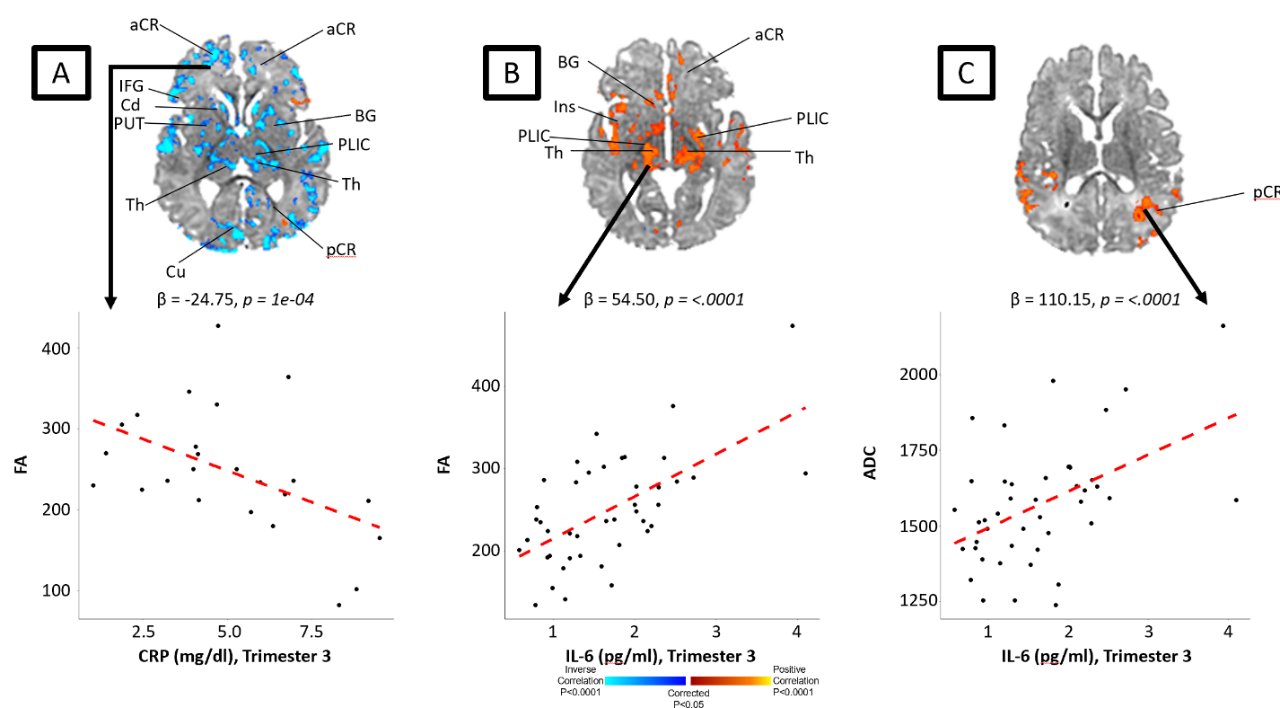

**Supplemental Figure 3:** **Associations between diffusion tensor imaging (DTI) fractional anisotropy (FA) and mean diffusivity (ADC), and maternal immune activation**. Parts A and B represent the association between DTI, FA, the dependent and maternal immune activation through IL-6 and CRP respectively during the 3^rd^ trimester as the independent, while part C represents the association between diffusion tensor imaging, ADC, the dependent and maternal immune activation through IL-6 during the 3^rd^ trimester as the independent. The red/yellow (positive) and purple/blue (inverse) areas show locations where maternal immune markers are associated with FA (images A,B) and ADC (image C). ALIC indicates anterior limb internal capsule; aCR, anterior region of corona radiata; BG, basal ganglia; Cd, caudate; CG, cingulum; CR: corona radiata; CC, corpus callosum; Cu, cuneus; IFO, inferior fronto-occipital; ITG, inferior temporal gyrus; Ins, Insula; MFG, medial frontal gyrus; MB, midbrain; MTG, middle temporal gyrus; pCR, posterior region of corona radiata; PUT, putamen; ST, stria terminalis; SFG, superior frontal gyrus; SFO, superior fronto-occipital; SLF, superior longitudinal fasciculus; SCR, superior region of corona radiata; STG, superior temporal gyrus; Th, thalamus; PLIC: posterior limb internal capsule.

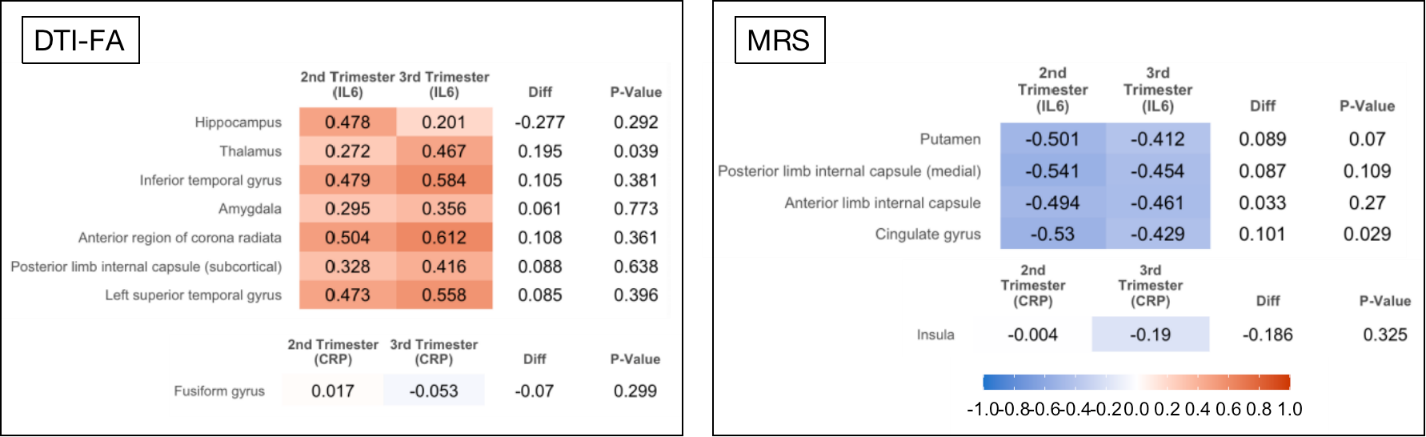

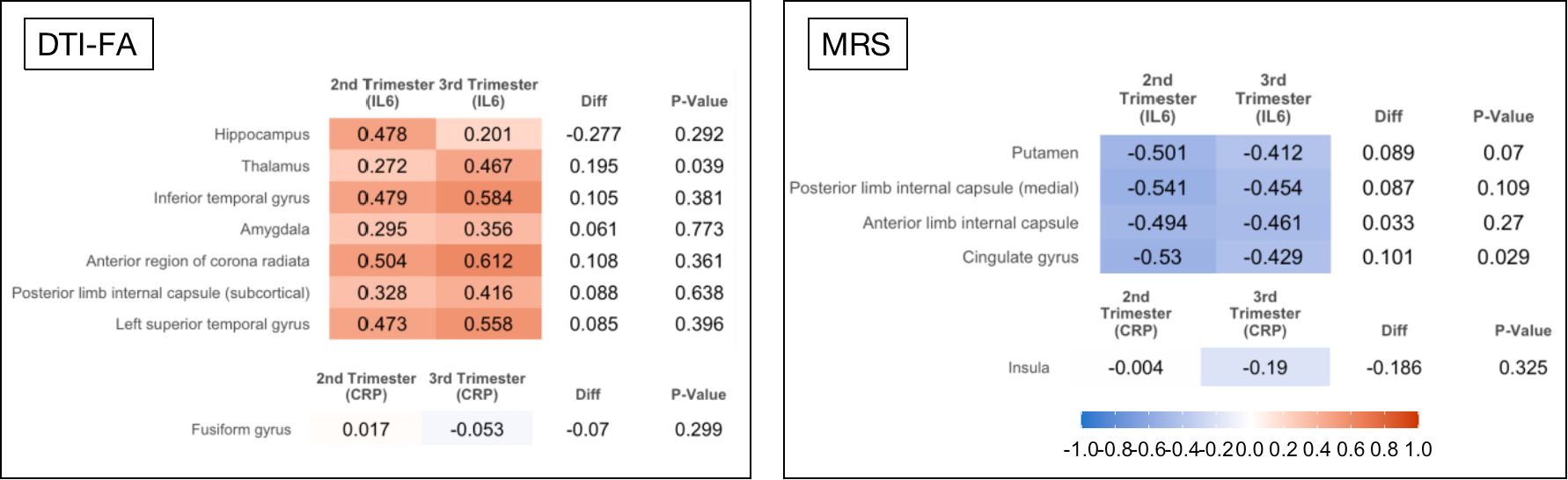

-1.0	-0.8	-0.6	-0.4	-0.2	0	0.2	0.4	0.6	0.8	1.0

**Supplemental Figure 4**: **Correlation comparisons between second and third trimester.** Comparing the strength of significance in the 2nd versus 3rd trimester for the regions of interest that were significantly associated with both immune markers. The correlation coefficients and their difference between the two trimesters are shown.
